# An individual-level socioeconomic measure for assessing algorithmic bias in health care settings: A case for HOUSES index

**DOI:** 10.1101/2021.08.10.21261833

**Authors:** Young J. Juhn, Euijung Ryu, Chung-Il Wi, Katherine S. King, Santiago Romero Brufau, Chunhua Weng, Sunghwan Sohn, Richard Sharp, John D. Halamka

## Abstract

While artificial intelligence (AI) algorithms hold great potential for improving health and reducing health disparities, biased AI algorithms have a potential to negatively impact the health of under-resourced communities or racial/ethnic minority populations. Our study highlights the major role of socioeconomic status (SES) in AI algorithm bias and (in)completeness of electronic health records (EHRs) data, which is commonly used for algorithm development. Understanding the extent to which SES impacts algorithmic bias and its pathways through which SES operates its impact on algorithmic bias such as differential (in)completeness of EHRs will be important for assessing and mitigating algorithmic bias. Despite its importance, the role of SES in the AI fairness science literature is currently under-recognized and under-studied, largely because objective and scalable individual-level SES measures are frequently unavailable in commonly used data sources such as EHRs. We addressed this challenge by applying a validated individual-level socioeconomic measure that we call the HOUSES index. This tool allows AI researchers to assess algorithmic bias due to SES. Although our study used a cohort with a relatively small sample size, these study results highlight a novel conceptual strategy for quantifying AI bias by SES.

## Introduction

Augmented computing power, storage capability and predictive analytics have accelerated the adoption and deployment of artificial (augmented or machine) intelligence (AI) tools in health care system of the United States (US).^1,2^ As of 2017, 96% of U.S. hospitals adopted certified EHRs and 98% of U.S. hospitals demonstrated meaningful use of at least one certified health information technology (HIT).^1^ A recent survey showed that 90% of health care executives in the US reported they had AI tools and automation strategy in 2020 compared to 53% in 2019.^2^ As of 2020, more than 80 imaging-related AI algorithms have been cleared by the US Food and Drug Administration (FDA).^3,4^ A research group at Mayo Clinic showed that an electrocardiogram (ECG)-based, AI-powered clinical decision support (CDS) tool demonstrated early diagnosis of low ejection fraction, a condition that is underdiagnosed but treatable, via a randomized clinical trial (RCT).^5^ The intervention increased the diagnosis of low ejection fraction in the overall cohort (1.6% in the control arm versus 2.1% in the intervention arm, odds ratio (OR) 1.32 (1.01– 1.61), P = 0.007).

While AI tools hold a great potential for improving health and even reducing health disparities, biased AI algorithm can negatively impact health in under-resourced or racial/ethnic minority populations as reported in the literature.^6-9^ For example, a recent study of a widely used commercial AI algorithm identified 18% of Black patients needing additional care for chronic disease management (compared to White patients). When a mitigation strategy was applied to reduce algorithmic bias, the percentage increased to 47% for Black patients^6^, demonstrating the potential harm of AI tools when the bias is not mitigated before implementation. Others reported algorithmic biases by race and socioeconomic status (SES) in post-partum depression,^10^ ICU mortality^11^ and 30-day psychiatric readmission.^11^ Therefore, while differential predictability of models by patient characteristics such as SES is a hardly new phenomenon in biomedical research,^12^ it has a major implication on health equity because of the potential to exacerbate inequities on a monumental scale when deployed on a large scale in clinical care.

Given the significant impact of SES on health risk and healthcare access, quantifying the degree of algorithmic bias by SES has important ethical implications for AI research as well as health care delivery research. SES is a key element of social determinants of health (SDH) in health care delivery and research^13-22^ and considered as a major factor accounting for differential health outcomes through their associated biological (eg, epigenetics, gene expression or telomere length), behavioral and environmental factors.^22-25^ Through influences on health care utilization, SES may shape the completeness of EHRs as a data source for developing AI algorithms and result in algorithmic bias by SES. Thus, SES is an important variable for understanding the nature of algorithmic bias stemming from differential health risk, healthcare access and completeness of EHRs and for addressing algorithmic bias in health care.

Unfortunately, individual-level SES measures that are objective, precise and scalable are frequently unavailable in commonly used data sources for clinical care and research^26^ posing a major barrier to health care delivery and research as acknowledged by National Academy of Medicine and National Quality Forum.^8,27,28^ Also, the limited availability of suitable individual-level SES in EHRs is a major challenge in addressing algorithmic bias in health care settings. Consequently, current AI fairness science focuses on readily available limited demographic factors such as age, sex and race/ethnicity,^29^ leaving the role of SES in AI bias poorly understood.

To address this major roadblock in the equitable implementation of health care AI, we propose to use the HOUSES (individual HOUsing-based SES) index to assess key features of SES in AI research such as accuracy, precision, objectivity and scalability. In this work, we 1) assessed differential data availability and validity of EHRs among study subjects according to SES as measured by HOUSES and 2) applied HOUSES index to quantify algorithmic bias by SES based on commonly used metrics for assessing AI bias.

## Methods

### Study Population and Setting

The study was conducted in primary care practices (i.e., including teaching pediatric faculty, residents, and nurse practitioners) in Olmsted County in southeastern Minnesota (MN). The study population and setting were described in our recent report.^30^ Briefly, Olmsted County, is a virtually self-contained health care environment with only two health care systems provide clinical care to nearly all residents. About 98% of residents authorize use of their medical records for research.^31^ According to U.S. census data in 2010, the age, sex, and ethnic characteristics of Olmsted County residents were similar to those of the state of Minnesota and the Upper Midwest.^32^ However, Olmsted County has become more diverse as indicated by the racial/ethnic characteristics of children enrolled in public schools (in 2019, 35.2 % reported to be a racial/ethnic minority groups). Mayo Clinic Primary Care Pediatric Practices offers primary care service at four locations within Olmsted County and this study was conducted in Baldwin Primary Care Practice site, the largest of the four practice sites. Asthma is the most prevalent chronic illness with the third highest health care expenditures in children and adolescents in Olmsted County, MN.^33^ The asthma prevalence in the primary care practice (14%) is slightly lower than that of the county (17.6%).^34^

### Study Design and Subjects

The study design and subjects were described in our recent report.^30^ Briefly, the study was designed as a cross-sectional study. We used the data from the training and testing cohorts that were used to develop ML algorithms applied to a previous study that developed a novel AI-assisted clinical decision support system named A-GPS (Asthma-Guidance and Prediction System), which is based on a single-center pragmatic randomized clinical trial (RCT). A-GPS-based intervention in the original study provided clinicians with the summary of most relevant information for asthma management and the risk prediction for asthma exacerbation using ML algorithms.^30^ It significantly reduced clinicians’ burden for EHRs review resulting in more efficient asthma management by providing more actionable guidance. The focus of the original study was to assess the effectiveness of A-GPS on asthma outcomes (e.g., asthma exacerbation, asthma control, asthma-related health care utilization, asthma care quality and health care costs). The original study used data from subjects who had persistent asthma or met Predetermined Asthma Criteria (PAC). In this present report, we limited the primary analysis assessing algorithmic bias to those with persistent asthma in order to focus on more homogeneous patient groups with persistent asthma. Details of the original study have been reported.^30^ For an additional analysis, we used the subjects in the original study who met PAC definition but were not yet diagnosed with asthma at the time of enrollment. This study (IRB number:15-004435) was approved by the Mayo Clinic Institutional Review Board (IRB).

### ML algorithms for predicting AE

For A-GPS, we trained and tested two ML algorithms: Naïve Bayes (NB) model and gradient boosting machine (GBM) model for predicting 1-year AE risk among children with asthma. We extracted 29 candidate variables based on the literature including sociodemographics, risk factors, and asthma outcomes from EHR over a prior three -year period. The original study included 590 subjects (300 in the training and 290 in the test set) who had persistent asthma or met Predetermined Asthma Criteria from Mayo Clinic pediatric practice panel, respectively. Receiver operating characteristic (ROC)-Areas Under Curve for NB and GBM model were 0.78 and 0.74 on the testing cohort, respectively. These algorithms were used to quantify algorithmic bias by SES in this report.

### Algorithmic fairness metrics

We used the widely reported common metrics as defined in Table 1. As it is impossible to satisfy all metrics (‘impossibility theorem’) ^35,36^ and the literature suggests to use false positive rate (FPR) and false negative rate (FNR),^35,36^ we used error rate, defined as the sum of FPR and FNR, as the primary metric for assessing algorithmic bias in this presented work. For each metric, we calculated the ratio comparing least privileged group (e.g., HOUSES Q1, see below) with the privileged group (HOUSES Q2-Q4). For FPR and error rate, ratio greater than 1 means that the algorithm is more favorable to privileged group, while more favorable to less privileged groups for the other three metrics (accuracy equality, equal opportunity and predictive parity). As a rule of thumb, a ratio that is less than 0.8 or greater than 1.25 (1/0.8) is considered as meaningful difference, which is implemented in an open source program such as AIF360.^37^

**Table 1.** Metrics for assessing algorithmic fairness used in this study

| Metric* | Definition** |
| --- | --- |
| Accuracy equality | $(TP+TN)/(TP+FP+TN+FN)$ |
| Equal opportunity (sensitivity) | $TP/(TP+FN)$ |
| Predictive equality (false positive rate) | $FP/(FP+TN)$ |
| Predictive parity (positive predictive value) | $TP/(TP+FP)$ |
| Error rate (FPR+FNR) | $FP/(FP+TN) + FN/(TP+FN)$ |
| * For each metric, ratio was by scores from unprivileged group, divided by that from privileged group. For FPR and error rate, with the ratio greater than 1 meaning that the algorithm is more favorable to privileged group, while more favorable to unprivileged group for the other three metrics.<br>** TP: true positives; FP: false positives; TN: true negatives; FN: false negatives; FPR: false positive rate; FNR: false negative rate |  |

Q1, see below) with the privileged group (HOUSESES Q2-Q4). For FPR and error rate, ratio greater than 1 means that the algorithm is more favorable to privileged group, while more favorable to less privileged groups for the other three metrics (accuracy equality, equal opportunity and predictive parity). As a rule of thumb, a ratio that is less than 0.8 or greater than 1.25 (1/0.8) is considered as meaningful difference, which is implemented in an open source program such as AIF360.<sup>37</sup>
HOUSESES: HOUSESES is an individual-level SES measure based on 4 real property data variables of an individual housing unit after principal component factor analysis: *housing value, square footage, number of bedrooms, and number of bathrooms*. An individual’s address from the EHR

### HOUSES

HOUSES is an individual-level SES measure based on 4 real property data variables of an individual housing unit after principal component factor analysis: *housing value, square footage, number of bedrooms, and number of bathrooms*. An individual’s address from the EHR is directly linked to the publicly available assessor’s data (which is a basis for property tax and thus is availale throughout US counties and cities). ^26^ We formulated a standardized HOUSES index score by summing these variables after z-score transformation. The greater the HOUSES index, the higher the SES. Since its development, HOUSES has been extensively applied as a validated SES measure that has shown association with numerous health-related outcomes, including acute/chronic conditions, healthcare access issues, healthcare utilization, and other health-related behaviors such as smoking and vaccine status as summarized in Table 2.

**Table 2.** The reported health outcomes predicted by HOUSES

|  |
| --- |
| **A. Chronic conditions and mortality** |
| **Adults:** 1) Risk of rheumatoid arthritis (RA) and post-RA mortality,<sup>38</sup> 2) risks of coronary heart disease, asthma, diabetes, hypertension, mood disorder,<sup>39</sup> Post-Myocardial Infarction mortality,<sup>40</sup> 3) multiple chronic conditions,<sup>41</sup> 4) post-Glioma mortality<sup>42</sup>, 5) renal transplantation outcome<sup>43</sup>, 6) ICU mortality,<sup>44</sup> 7) persistent depressive symptoms and remission of depressive symptoms,<sup>45</sup> |
| **Children:** 8) Risks of asthma, epilepsy, mood disorders,<sup>46</sup> 9) overweight,<sup>26,47</sup> 8) asthma outcome<sup>48</sup>, 10) multiple complex chronic conditions,<sup>49</sup> 11) serum 25 (OH)D concentration<sup>50</sup> |
| **B. Acute conditions** |
| **Adults:** 12) Risk of accidental falls<sup>51</sup>, 13) osteoporotic fracture incidence<sup>52</sup> |
| **Children:** 14) Risk of invasive pneumococcal disease,<sup>53</sup> 15) risk of bronchiolitis, pneumonia, urinary tract infection, adverse childhood experiences<sup>46</sup> |
| **C. Other health outcomes** |
| **Adults:** 16) All-cause hospitalization,<sup>41</sup> 17) smoking status,<sup>54</sup> 18) detecting survey biases,<sup>55</sup> 19) end of life care access (advance directives, social work consultation)<sup>56</sup> |
| **Children:** 20) Low birth weight, household smoking status,<sup>26,47</sup> 21) pertussis vaccine up-to-date status,<sup>57</sup> 22) self-rated health<sup>58</sup>, 23) HPV vaccine (initiation and completion) up-to-date status<sup>59</sup> |

### Other pertinent variables

While our focus is to quantify algorithmic bias by SES, we also considered other readily available demographic characteristics (age, sex, and race/ethnicity), and pediatric chronic conditions defined by Feudtner et al (an accepted measure of pediatric chronic conditions in literature).^49^ These variables are extracted from patient’s EHR. For chronic conditions, ICD-9 diagnostic and procedure codes were used. For simplicity, age (<12 vs ≥12 years) and chronic conditions (yes vs no) are dichotomized. To demonstrate the impact of SES on completeness of EHR, we compared availability of 7 key variables that are clinically relevant to childhood asthma management (health maintenance visit, asthma compliance, asthma severity, asthma type, NAEPP recommendation, smoking status, and missing school; See Table 2). These variables were extracted from EHR in the 3 years prior to the study index date. Additionally, we assessed data validity defined as having ICD-9 codes for asthma among those who met PAC definition but were not yet diagnosed with asthma at the time of the study.^30^ Specifically, we previously reported a significant number of children had undiagnosed asthma by comparing asthma prevalence by ICD code-based asthma ascertainment with that by NLP-based ascertainment using PAC (sensitivity 31% (ICD-9) vs. 81% (NLP logic) and 85% (NLP ML)).^60-62^

### Data analysis

In this presented work, we quantified algorithmic bias for two ML algorithms (NB and GBM models for predicting 1-year AE risk among pediatric asthmatics) by demographic factors (age, sex, race/ethnicity), SES (HOUSES), and chronic condition. This was done using the testing cohort, because model performance metrics in the training cohort are generally overestimated. To see the impact of SES on completeness of EHR, we also calculated proportions of subjects with missing or unknown information for 7 key variables for asthma management measuring data availability. Based on our earlier work, we assessed one variable measuring data validity (i.e., diagnosed vs. undiagnosed asthma by ICD codes for those who met PAC^61,62^) by SES. This calculation was done in both the training and testing cohorts.

## Results

### Subject characteristics

Subjects included in the training cohort were younger (71% with younger than 12 years old), more male (57%), more non-Hispanic White (57%)) as shown in Table 3. Roughly 20% of the subjects were in the low-SES (HOUSES Q1) group and 20 % had at least one chronic condition. Subject characteristics were similar between training and testing cohorts. Roughly 30% of subjects had asthma exacerbation within 1-year follow-up period (26% in the training cohort and 35% in the testing cohort: Table 3). Table 4 showed that proportion of AE differed by subject characteristics. In general, the proportion was higher in subjects who were younger, male, lower SES, and those with chronic conditions.

**Table 3.** Subject characteristics used in the study

| **Table 3.** Subject characteristics used in the study |  |  |
| --- | --- | --- |
|  | Training cohort<br>(N=133) | Testing cohort<br>(N=113) |
| Age (in years), n (%) |  |  |
| < 12 | 94 (71%) | 80 (71%) |
| ≥ 12 | 39 (29%) | 33 (29%) |
| Sex, n (%) |  |  |
| Male | 76 (57%) | 67 (59%) |
| Female | 57 (43%) | 46 (41%) |
| Race/ethnicity, n (%) |  |  |
| Non-Hispanic Whites | 76 (57%) | 67 (59%) |
| Others | 57 (43%) | 46 (41%) |
| HOUSES, n (%) |  |  |
| Q1 (the lowest SES) | 22 (18%) | 15 (14%) |
| Q2-Q4 | 102 (82%) | 92 (86%) |
| Missing | 9 | 6 |
| Chronic condition, n (%) |  |  |
| Yes | 30 (23%) | 19 (17%) |
| No | 103 (77%) | 94 (83%) |
| Asthma exacerbation, n (%) |  |  |
| Yes | 34 (26%) | 40 (35%) |
| No | 99 (74%) | 73 (65%) |

**Table 4.** Proportion of subjects with asthma exacerbation (AE) by subject characteristics

|  |  |  |
| --- | --- | --- |
|  | Percentage of subjects with AE |  |
|  | Training cohort | Testing cohort |
| Age (in years), n (%) |  |  |
| < 12 | 28 (29.8%) | 30 (37.5%) |
| ≥ 12 | 6 (15.4%) | 10 (30.3%) |
| Sex, n (%) |  |  |
| Male | 25 (32.9%) | 23 (34.3%) |
| Female | 9 (15.8%) | 17 (37.0%) |
| Race/ethnicity, n (%) |  |  |
| Non-Hispanic Whites | 19 (25.0%) | 25 (37.3%) |
| Others | 15 (26.3%) | 15 (32.6%) |
| HOUSES, n (%) |  |  |
| Q1 (the lowest SES) | 6 (27.3%) | 8 (53.3%) |
| Q2-Q4 | 23 (22.5%) | 29 (31.5%) |
| Chronic condition, n (%) |  |  |
| Yes | 10 (33.3%) | 7 (36.8%) |
| No | 24 (23.3%) | 33 (35.1%) |

### Algorithmic bias

Using the testing cohort, Table 5 summarizes the results of algorithmic bias for both NB and GBM models for predicting 1-year AE risk. Overall, algorithmic performance depended on patient characteristics such as age, sex, and chronic diseases as expected. Also, the degree of differential algorithmic performance by these factors depends on ML algorithm used (NB vs GBM model) in a way that no specific ML algorithm was more or less susceptible to algorithmic bias.

**Table 5.**
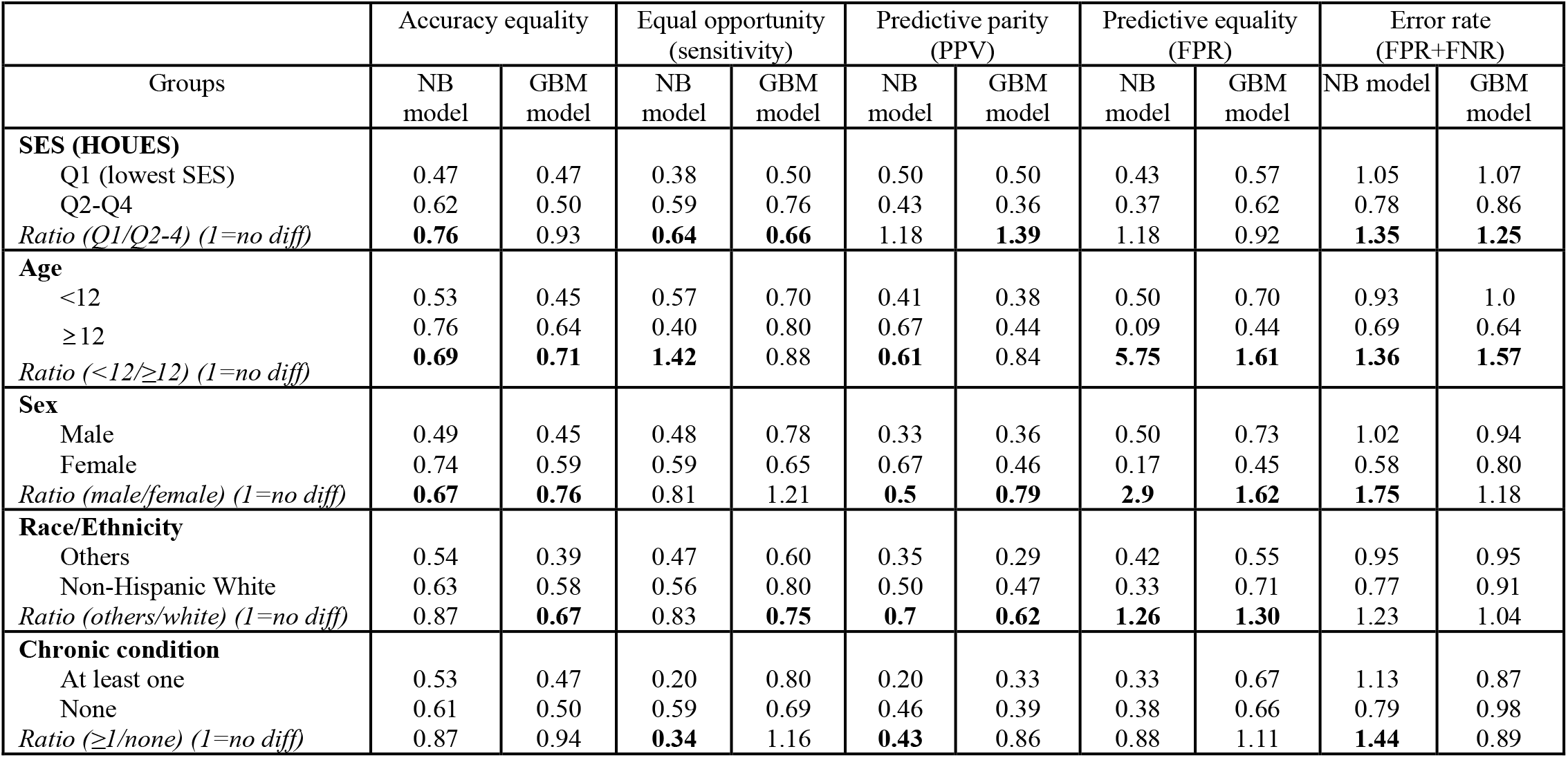
Assessment of algorithmic bias for two machine learning algorithms (Naïve Bayes [NB] and gradient boosting machine [GBM] models) predicting 1-year asthma exacerbation risk in childhood asthma using 5 commonly used bias metrics.

|  |  |  |  |  |  |  |  |  |  |  |
| --- | --- | --- | --- | --- | --- | --- | --- | --- | --- | --- |
|  | Accuracy equality |  | Equal opportunity (sensitivity) |  | Predictive parity (PPV) |  | Predictive equality (FPR) |  | Error rate (FPR+FNR) |  |
| Groups | NB model | GBM model | NB model | GBM model | NB model | GBM model | NB model | GBM model | NB model | GBM model |
| **SES (HOUES)** |  |  |  |  |  |  |  |  |  |  |
| Q1 (lowest SES) | 0.47 | 0.47 | 0.38 | 0.50 | 0.50 | 0.50 | 0.43 | 0.57 | 1.05 | 1.07 |
| Q2-Q4 | 0.62 | 0.50 | 0.59 | 0.76 | 0.43 | 0.36 | 0.37 | 0.62 | 0.78 | 0.86 |
| *Ratio (Q1/Q2-4) (1=no diff)* | **0.76** | 0.93 | **0.64** | **0.66** | 1.18 | **1.39** | 1.18 | 0.92 | **1.35** | **1.25** |
| **Age** |  |  |  |  |  |  |  |  |  |  |
| <12 | 0.53 | 0.45 | 0.57 | 0.70 | 0.41 | 0.38 | 0.50 | 0.70 | 0.93 | 1.0 |
| ≥12 | 0.76 | 0.64 | 0.40 | 0.80 | 0.67 | 0.44 | 0.09 | 0.44 | 0.69 | 0.64 |
| *Ratio (<12/≥12) (1=no diff)* | **0.69** | **0.71** | **1.42** | 0.88 | **0.61** | 0.84 | **5.75** | **1.61** | **1.36** | **1.57** |
| **Sex** |  |  |  |  |  |  |  |  |  |  |
| Male | 0.49 | 0.45 | 0.48 | 0.78 | 0.33 | 0.36 | 0.50 | 0.73 | 1.02 | 0.94 |
| Female | 0.74 | 0.59 | 0.59 | 0.65 | 0.67 | 0.46 | 0.17 | 0.45 | 0.58 | 0.80 |
| *Ratio (male/female) (1=no diff)* | **0.67** | **0.76** | 0.81 | 1.21 | **0.5** | **0.79** | **2.9** | **1.62** | **1.75** | 1.18 |
| **Race/Ethnicity** |  |  |  |  |  |  |  |  |  |  |
| Others | 0.54 | 0.39 | 0.47 | 0.60 | 0.35 | 0.29 | 0.42 | 0.55 | 0.95 | 0.95 |
| Non-Hispanic White | 0.63 | 0.58 | 0.56 | 0.80 | 0.50 | 0.47 | 0.33 | 0.71 | 0.77 | 0.91 |
| *Ratio (others/white) (1=no diff)* | 0.87 | **0.67** | 0.83 | **0.75** | **0.7** | **0.62** | **1.26** | **1.30** | 1.23 | 1.04 |
| **Chronic condition** |  |  |  |  |  |  |  |  |  |  |
| At least one | 0.53 | 0.47 | 0.20 | 0.80 | 0.20 | 0.33 | 0.33 | 0.67 | 1.13 | 0.87 |
| None | 0.61 | 0.50 | 0.59 | 0.69 | 0.46 | 0.39 | 0.38 | 0.66 | 0.79 | 0.98 |
| *Ratio (≥1/none) (1=no diff)* | 0.87 | 0.94 | **0.34** | 1.16 | **0.43** | 0.86 | 0.88 | 1.11 | **1.44** | 0.89 |

SES as measured by HOUSES index *greatly* impacted algorithmic performance. Specifically, children in lower SES groups had higher error rates than those in the higher SES group in both ML models (ratio = 1.35 for NB model and 1.25 for GBM model) which exceed those for race/ethnicity (1.23 and 1.04, respectively). This differential algorithmic bias by SES was driven more by FNR (=1-sensitivity; ratio=1.51 by NB and 2.01 by GBM model) than FPR (1.18 by NB and 0.92 by GBM model). This was also true for equal opportunity (i.e., sensitivity) metric. Children in the higher SES group had significantly higher sensitivity of both algorithms, compared to those in the lower SES group in a way exceeding the impact of other demographic factors.

### Availability and accuracy of data relevant to asthma management

We compared data availability for the key variables associated with the risk of AE in the training and testing cohorts. As shown in Table 6, compared to children in the higher SES group, those from lower SES background had higher unavailability of the key variables for asthma (eg, compliance data, severity and smoking exposure) associated with the risk of AE. Additionally, children with lower SES had higher prevalence of undiagnosed asthma, compared to those with higher SES, although they met the criteria for asthma.

**Table 6:** Summary of data availability for variables relevant to asthma management and data validity by SES for each cohort (training and testing cohort)

|  | Training |  | Testing |  |
| --- | --- | --- | --- | --- |
| **Data unavailability, n (%)** | Q1 (n=22) | Q2-Q4 (n=102) | Q1 (n=15) | Q2-Q4 (92) |
| Missing health maintenance visit | 3 (14%) | 12 (12%) | 2 (13%) | 11 (12%) |
| Missing asthma compliance | 14 (64%) | 43 (42%) | 9 (60%) | 44 (48%) |
| Missing asthma severity | 9 (41%) | 24 (24%) | 8 (53%) | 23 (25%) |
| Missing asthma type | 22 (100%) | 95 (93%) | 14 (93%) | 79 (86%) |
| NAEPP recommendation missing | 13 (59%) | 43 (42%) | 8 (53%) | 37 (40%) |
| Missing smoking status | 16 (73%) | 39 (38%) | 8 (53%) | 34 (37%) |
| Missing data on missing school | 15 (68%) | 41 (40%) | 8 (53%) | 34 (37%) |
|  | Training |  | Testing |  |
| **Data validity\*** | Q1 (n=34) | Q2-Q4 (n=112) | Q1 (n=37) | Q2-Q4 (n=121) |
| Undiagnosed (ICD) asthma | 4 (12%) | 11 (9.8%) | 3 (8.1%) | 8 (6.6%) |

\* Data validity was calculated for subjects who met PAC criteria but did not have physician diagnosis of asthma

## Discussion

Our study results suggest that SES as measured by HOUSES may impact algorithmic bias in a way resulting in greater algorithmic bias in patients with lower SES, compared to those with higher SES. Additionally, children with asthma from lower SES groups had a greater degree of unavailable and inaccurate EHRs data for asthma management, compared to those with higher SES. One noteworthy finding is disparities in undiagnosed or delayed diagnosed asthma by SES as the lack of timely diagnosis of asthma will deter access to preventive and therapeutic interventions^63,64^ and may impact long-term respiratory outcomes. We postulate SES might impact algorithmic bias through differential completeness of EHRs data as SES impacts health risk, healthcare access and completeness of EHRs.

As discussed earlier, SES is a key variable for understanding the nature of algorithmic bias stemming from differential health risk, health care access, and completeness of available EHRs and for assessing and mitigating algorithmic bias in health care. However, objective, scalable and well-validated individual-level SES measures are frequently unavailable in commonly used data sources for clinical care and research^26^ posing a major barrier to health care delivery and research as acknowledged by National Academy of Medicine and National Quality Forum.^8,27,28^ In this respect, HOUSES measuring individual-level SES, can be a useful tool for health care research including AI research as it overcomes such unavailability of individual-level SES measures in commonly used data sources such as EHRs.

Our previous work demonstrated that SES defined by HOUSES index predicted a broad range of health outcomes and care quality as summarized in Table 2. Relevant to this present report, we showed that HOUSES was associated with inconsistent self-reporting.^55^ We found that lower HOUSES (SES) was associated with higher rates of inconsistency (inaccuracy) in -self-reporting a diagnosed disease for the given (documented) diseases between the baseline and 4-year follow-up survey, and the association remained significant after pertinent characteristics such as age and perceived general health (adjusted OR=1.46; 95% CI 1.17 to 1.84 for the lowest compared with the highest HOUSES decile). Given that self-reported information is captured in EHR and often used clinically (e.g., a history of pediatric asthma), higher proportion of inconsistent self-reporting among patients with low SES may produce less reliable ML algorithms (if used). In addition, our unpublished data showed that availability of patient’s online portal system (a proxy for healthcare access) was significantly lower among families with lower SES (68% in Q1 [lowest SES]), compared to 74% in Q2, 88% in Q3, and 92% in Q4 (highest SES) (p=.02). As online portal is an important tool for chronic disease management such as childhood asthma (eg, communicating with care providers, patient reported outcomes [PROs], medication update, etc, being captured in EHRs), it significantly affected availability of a key PROs on asthma (i.e., Asthma Control Test results; 99% for those with portal vs. 77% for those without portal) at the end of a clinical trial. Indeed, our study results in Table 5 showed the potential impact of SES as measured by HOUSES on algorithmic bias. For example, error rates were higher for children with lower SES for both algorithms predicting the risk of AE, compared to those with higher SES and exceed the impact of other demographic factors (age, sex and race/ethnicity). This was also true for sensitivity. A recent study also showed SES defined by health insurance (public vs. commercial health insurance) influenced ML algorithms predicting ICU mortality^11^ and 30-day psychiatric readmission (people with lower SES had poorer prediction performance of their ML algorithms, compared to those with higher SES).^11^ Overall, our study results and the literature suggest that SES impacts differential (in)completeness and validity of PROs which may influence differential algorithmic performance by SES if algorithms are trained by using skewed training cohort by individual-level SES.

It is also important to recognize differential performance of SES measures in predicting health outcomes because researchers routinely use aggregate-level SES measures ^18,65-67^or other SES measures in research. Our recent study showed that HOUSES predicted that kidney transplant recipients with lowest HOUSES (Q1) had a significantly higher risk of graft failure than those with highest HOUSES (Q2-4) (adjusted hazard ratio: 2.12; 95% CI, 1.08-4.16).^43^ Importantly, other SES measures such as individual educational levels and census-block group level education and income failed to predict outcomes on graft failure. Therefore, in assessing and mitigating algorithmic bias by SES, it is important to use an SES measure with accuracy and precision in capturing individual-level SES measure and in this respect, HOUSES fulfills these requirements and can be a complementary SES measure beyond the existing conventional SES measures. HOUSES has several conceptual and methodological merits for clinical and translational research: First, HOUSES is able to capture health effects of SES (defined as ‘one’s ability to access desired resources’)^68^ which predicts 39 health care access, care quality, and health outcomes as summarized in Table 2. Second, it assesses an objective individual-level SES measure, in contrast to self-reported (e.g., income) or aggregate-level (e.g., zip-code based Census data) measures. Third, it can retrospectively measure SES at any given point in time whenever address information at the index date of events is available (not relying on recalls). Fourth, as spatial coordinates are intrinsic to HOUSES, it enables geospatial analysis to identify geographic hotspots of interest (e.g., COVID-19 cases) to be used as a feature in predictive models.^69-71^ Finally, unlike other SES measures (e.g., educational level which is relatively static), it can capture longitudinal changes as real property data are regularly updated, and relocation of residence often reflects changes in a subject’s SES. This feature allows us to use HOUSES as a financial outcome across life stages. Taken together, these features highlight how the HOUSES measure can help to address issues of algorithmic fairness, ultimately helping to achieve greater levels of health equity across populations.

Our study has a few strengths. First, our study is based on a real world setting where patients have a wide range of EHR completeness, instead of studies based on highly selected subjects. Second, we used an objective individual-level SES measure instead of self-reported or aggregate-level SES measures (e.g., Census level data). Therefore, it does not suffer from biases such as recall bias or inaccuracy due to aggregation. Third, we assessed data availability and validity for features relevant to AE risk, which is not commonly done in AI research despite its importance. Our study also has limitations. First, the analysis was based on a small sample size. However, this pilot study provides a conceptual framework for using SES when assessing AI bias. Second, our study subjects may not represent the general pediatric population. However, it represents patient population (source population) as this study was based on those who receive care at Mayo Clinic without involving any recruitment steps.

In conclusion, our study findings highlight the important role of SES in assessing algorithmic bias. Understanding the extent to which SES impacts contributes to algorithmic bias, and examining the potential impact of SES on emerging applications of AI in healthcare will be crucially important for recognizing and mitigating algorithmic bias, ultimately supporting efforts to promote health equity and fairness. We believe the HOUSES index can play an important role in those efforts.

## Data Availability

The datasets generated and/or analyzed during the current study are not publicly available as they include protected health information. Access to data could be discussed per the institutional policy after approval of the IRB at Mayo Clinic.

## Acknowledgement

We would like to acknowledge the HOUSES program of the Mayo Clinic and Precision Population Science Lab staff, as well as thank Ms. Kelly Okeson for her administrative assistance.

## Abbreviations

AE: Asthma exacerbation
AI: Artificial Intelligence
EHRs: Electronic health records
FN: False negatives
FP: False positives
GBM: Gradient Boosting Machine
HOUSES: HOUsing-based SocioEconomic Status measure
ML: Machine Learning
NB: Naïve Bayes
NAEPP: National Asthma Education and Prevention Program;
PAC: Predetermined Asthma Criteria
SDH: Social Determinants of Health
SES: Socioeconomic status
TN: True negatives
TP: True positives

